# Efficacy of Mosquito Shield^TM^, a transfluthrin spatial emanator against wild, free-flying pyrethroid-resistant *Anopheles gambiae* s.l.; an experimental hut evaluation in Benin, West Africa

**DOI:** 10.64898/2026.02.19.26346635

**Authors:** Boris N’dombidjé, Thomas Syme, Juniace Ahoga, Benjamin Pearce, Ange Yadouleton, Melis Yamadjako, Corine Ngufor

**Author notes:** Correspondence: (CN). Email addresses: BN, TS, JN, BP, AY, MY, CN.

## Abstract

**Background:** Spatial emanators disrupt mosquito behaviour by inducing movement away from chemical stimuli and interfering with host detection and feeding. These tools were recently endorsed by the World Health Organization (WHO) for malaria control, based largely on clinical evidence from East Africa. Mosquito Shield^TM^ is a passive, transfluthrin-based emanator designed to provide month-long protection in enclosed or semi-enclosed spaces. This study evaluated its entomological efficacy under experimental hut conditions in Benin, West Africa to generate evidence in support of WHO prequalification.

**Methods:** An experimental hut trial was conducted against wild free-flying pyrethroid-resistant *Anopheles gambiae* s.l. at the Covè field station in southern Benin over two 32-day product life cycles of Mosquito Shield^TM^. Sixteen West African–style experimental huts were assigned to Mosquito Shield^TM^ or a placebo control. Efficacy was measured using human landing catches (HLC) and mosquito aspirations following standard hut testing methods. Primary endpoints included protective efficacy against mosquito landing (HLC) and personal protection against blood-feeding (mosquito aspiration). Secondary endpoints included deterrence, exophily, mortality, and blood-feeding inhibition. WHO susceptibility bioassays were conducted on the local *An gambiae s.l.* population to investigate susceptibility to public health insecticides during the trial.

**Results:** WHO susceptibility bioassays confirmed high levels of resistance to pyrethroids, including transfluthrin, in the local *Anopheles gambiae* s.l. population. A total of 5,682 *An. gambiae* s.l. and 6,158 *Mansonia africana* were collected through HLCs, and 1,436 *An. gambiae* s.l. by mosquito aspirations. Mosquito Shield^TM^ significantly reduced mosquito landing, providing 43.0% protective efficacy (95% CI: 24.0–57.0; *p* < 0.001) against *An. gambiae* s.l. and 38.0% protective efficacy (95% CI: 12.0–57.0; *p* = 0.008) against *Mansonia africana*. Mosquito aspiration data showed 48.5% deterrence, 29.9% blood-feeding inhibition, and 64% personal protection (95% CI: 21.9–81.8; *p* < 0.001) against *An. gambiae* s.l. No mosquito mortality was observed in the control huts, whereas mortality of *An. gambiae* s.l. reached 49.0% in HLC collections and 22.7% in aspiration collections. Mosquito Shield^TM^ also induced >96% mortality in *Mansonia africana*, demonstrating both lethal and behavioural effects against both vector species.

**Conclusions:** Mosquito Shield^TM^ significantly reduced mosquito entry, landing, blood-feeding, and survival of pyrethroid-resistant *An. gambiae* s.l. under semi-field experimental hut conditions in West Africa, with additional effects against *Mansonia africana*. These results support its WHO prequalification and highlight its potential as a complementary vector control tool to strengthen malaria prevention and provide additional benefits for integrated control of other vector-borne diseases.

## Introduction

Between 2000 and 2015, large-scale deployment of insecticide-treated nets (ITNs) and indoor residual spraying (IRS) contributed significantly to reductions in malaria burden worldwide [1]. These core vector control tools remain central to malaria prevention strategies, but their impact is increasingly under threat. In recent years, progress has stalled and risks reversing without the introduction of new interventions [2]. The stagnation is largely driven by the widespread emergence of insecticide resistance in malaria vectors, declining durability and household coverage of ITNs, reduced implementation and coverage of IRS, and diminishing financial investment in malaria control. Together, these challenges underscore the urgent need for complementary tools that can strengthen existing interventions, close protection gaps left by ITNs, and sustain progress toward malaria elimination.

Spatial emanators are airborne compounds that disrupt mosquito behaviour by inducing movement away from a chemical stimulus and interfering with host detection and feeding [3]. When volatilized into the air within a defined space, these compounds can reduce mosquito–human contact and thereby provide personal or household protection, ultimately lowering the risk of disease transmission [3–5]. Conventional commercial products, such as mosquito coils and electrical plug-ins, have long been used to provide this form of protection. However, these products rely on a heat source to volatilize the active ingredient (AI), which not only limits convenience and user compliance but also poses potential health risks from smoke exposure [6–8]. To address these limitations, passive emanator spatial repellents have been developed. These devices release volatile concentrations of repellent AI at room temperature through natural air movement, requiring little or no user action [9]. Their efficacy has been long demonstrated in reducing human–vector contact with Aedes mosquitoes, the primary vectors of arboviral diseases such as dengue, chikungunya, and Zika. More recently, based on clinical evidence from trials in Kenya [3], China [4] and Indonesia [5], the World Health Organization (WHO) has issued a conditional recommendation supporting their use indoors, in addition to ITNs, to prevent and control malaria in both children and adults in areas with ongoing transmission [6, 7].

Mosquito Shield^TM^ developed by SC Johnson & Son Inc. is a passive spatial emanator consisting of a multilayer plastic pouch impregnated with 110 mg of transfluthrin. When hung in enclosed or semi-enclosed spaces, the active ingredient is released into the surrounding air through natural airflow, providing continuous protection against mosquito bites for up to one month without the need for external energy sources. In a recent cluster randomized controlled trial (cRCT) in Kenya, Mosquito Shield^TM^ reduced malaria infections by about one-third, providing the first evidence of epidemiological impact of spatial emanators in Africa alongside high ITN coverage [3]. While evidence of its epidemiological impact in West Africa is yet to be demonstrated, a small scale household randomized trial conducted in Benin demonstrated a 34.2% reduction in human landing rates in an area of intense pyrethroid resistance [8]. For Mosquito Shield^TM^ to be eligible for procurement by United Nations agencies, it must first be included on the WHO list of prequalified vector control products. This process requires a comprehensive assessment of the product’s quality, safety, and efficacy, both under controlled conditions and across multiple settings relevant to its intended use.

To support its WHO prequalification assessment, we conducted a Good Laboratory Practice (GLP, OECD)–compliant experimental hut trial at the Covè station in Benin. The study evaluated the efficacy of Mosquito Shield^TM^ in preventing landing and blood-feeding of wild, free-flying, pyrethroid-resistant *Anopheles gambiae* s.l. in West African–type experimental huts.

## Materials and methods

### Study site

The experimental hut trial was conducted at a field station in Covè, southern Benin (7°14′N; 2°18′E). The station lies within an extensive rice irrigation zone that provides permanent breeding sites for mosquitoes. Both *Anopheles coluzzii* and *An. gambiae* s.s. occur sympatrically, with *An. coluzzii* predominating. Recent studies have documented high frequency and intensity of resistance to pyrethroids and organochlorines, while susceptibility to carbamates, organophosphates, and pyrroles is maintained [9]. Molecular analyses have shown that pyrethroid resistance is associated with a high prevalence of the knockdown resistance (kdr) L1014F mutation and overexpression of cytochrome P450 monooxygenases [10].

### Experimental hut trial

#### Experimental hut design and treatments

The trial was conducted in West African–style experimental huts at Covè. Each hut is built of concrete bricks with cement-plastered walls, a corrugated iron roof, and a cement ceiling. A wooden-framed veranda projects from the rear wall to capture mosquitoes exiting due to behavioural or insecticidal effects of treatments. The huts have a mean volume of 21.1 m³ and include a room compartment (2.1 × 3.4 × 2.4 m) and a veranda compartment (1.7 × 1.7 × 1.5 m). Entry of wild mosquitoes is enabled through four 1 cm–wide window slits located on the walls of each hut, while access by natural predators is prevented by water-filled moats constructed around the hut. In total, 16 huts were used. To avoid contamination from earlier trials, all walls were replastered and allowed to cure for one month before the study commenced. Eight huts were randomly assigned to the Mosquito Shield^TM^ treatment arm and eight to the placebo control arm.

Four Mosquito Shield^TM^ devices were installed per hut based on the size of the experimental huts. They were affixed to the walls with nails and wooden supports at approximately two-thirds of wall height, in accordance with the manufacturer’s instructions (Figure 1). The trial was conducted over two consecutive 32-day product life cycles, with new Mosquito Shield^TM^ devices applied at the start of each round. The first round took place July 10^th^ to August 10^th^ 2024 and the second November 11^th^ to December 10^th^ 2024, allowing data collection across different transmission seasons.

**Figure 1.**
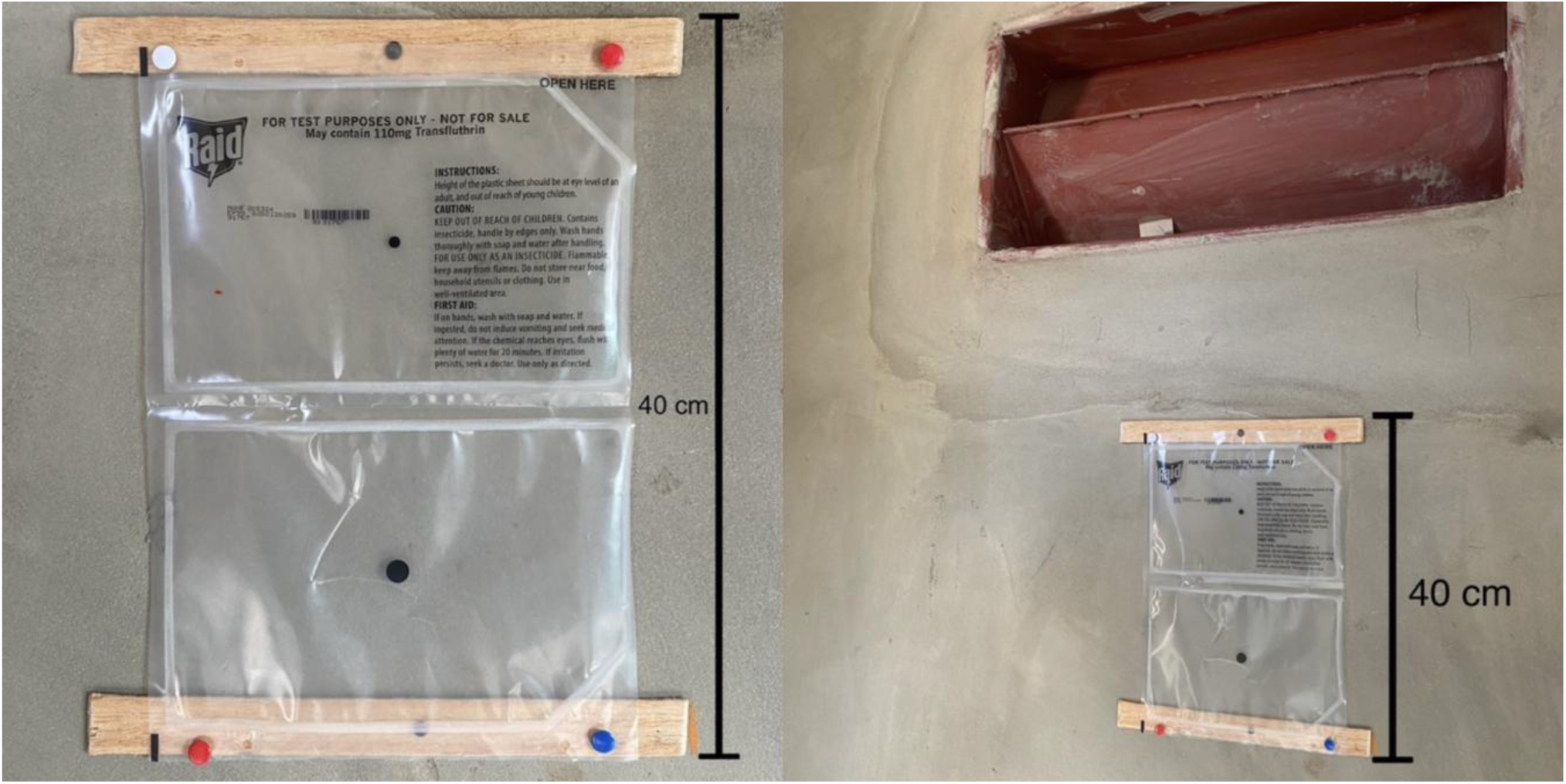
Installation of Mosquito Shield^TM^ devices on the interior walls of experimental huts, affixed with nails and wooden supports at approximately two-thirds of wall height.

#### Mosquito collection methods

During each 32-day trial round, mosquito collections were conducted in four replicate huts per treatment using human landing catches (HLC) and in the other four huts using aspiration. To minimize positional bias, collection methods were rotated between hut sets every four days, ensuring that by the end of each round, each method was applied for 16 nights per hut.

***Human Landing Catches (HLCs):*** HLCs were conducted by trained volunteers who collected mosquitoes landing on their legs throughout the night to estimate human landing rates. Sixteen consenting collectors were recruited per round and organized into eight pairs, rotated across replicate huts on successive nights to reduce bias due to individual attractiveness to mosquitoes. Each pair alternated shifts, with one collector working from 19:00–01:00 and the other from 01:00–07:00.

***Mosquito aspiration method:*** This method followed standard WHO experimental hut trial procedures [11] typically used for evaluation of indoor vector control tools. A volunteer sleeper occupied each hut from 21:00 to 06:00, protected by an untreated bed net to reduce biting pressures. Each net had six holes (4 × 4 cm) in accordance with WHO ITN evaluation guidelines to allow mosquito entry and assessment of blood-feeding protection. Each morning at 06:00, mosquitoes were collected from under the net, room and veranda compartments using a torch and mouth aspirator, placed in labelled plastic cups, and transported to the laboratory for assessment of mortality, blood-feeding inhibition, deterrence, and exophily. To control for variation in individual attractiveness to mosquitoes, sleepers were rotated between huts on successive nights.

#### Experimental hut trial outcome measures

The efficacy of Mosquito Shield^TM^ in the experimental huts, was measured using the following primary and secondary endpoints for each collection method:

##### Primary endpoints for HLC method

The primary endpoint for the HLC method was protective efficacy that was calculated both crudely and using model-adjusted estimates.

- Crude protective efficacy was measured as a direct reduction in the number of mosquitoes collected in Mosquito Shield––treated huts compared with control huts as follows:

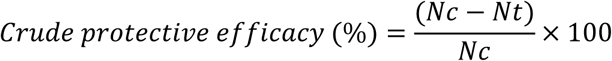

Where ‘Nc’ is the number of mosquitoes collected in the control arm and ‘Nt’ is the number collected of mosquitoes collected in the treated arm.
- Model-adjusted protective efficacy was calculated as follows:

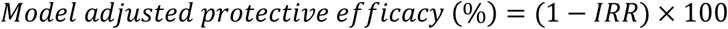

Where ‘IRR’ is the model-derived incidence rate ratio describing the difference in the number of mosquitoes collected in treated huts relative to the control.

##### Primary endpoints for mosquito aspiration method

The primary endpoint for the mosquito aspiration method was personal protection, also calculated crudely and using model adjusted estimates:

- Crude personal protection was calculated as a direct reduction in the number of blood-fed mosquitoes collected in Mosquito Shield––treated huts compared with control huts as follows:

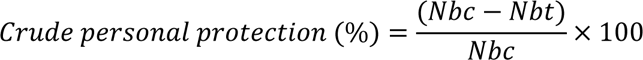

Where ‘Nbc’ is the number of blood-fed mosquitoes collected in the control huts and ‘Nbt’ is the number collected of blood-fed mosquitoes collected in the Mosquito Shield^TM^ huts.
- Model-adjusted personal protection was calculated as follows:

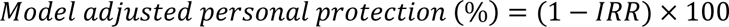

Where ‘IRR’ is the model-derived incidence rate ratio describing the difference in the number of blood-fed mosquitoes collected in treated huts relative to the control.

##### Secondary endpoints

*Human Landing Catches (HLC):*

- Mosquito mortality: The proportion of mosquitoes found dead 24 hours after collection

*Mosquito aspirations:*

- Mosquito mortality: The proportion of mosquitoes found dead 24 hours after collection
- Deterrence: Reduction in hut entry rates compared to the control.
- Exophily: Proportion of mosquitoes collected in the veranda, indicating the tendency to exit the hut.
- Proportion inside net: Percentage of mosquitoes collected under the untreated mosquito net.
- Blood-feeding rate: Proportion of blood-fed mosquitoes collected per treatment.
- Blood-feeding inhibition: Reduction in the proportion of blood-fed mosquitoes in treated huts relative to controls.

### WHO susceptibility bioassays

WHO tube tests and bottle bioassays were conducted using F1 progeny of *Anopheles gambiae* s.l. larvae collected from breeding sites near the experimental hut station to assess vector susceptibility to transfluthrin and other commonly used malaria control insecticides during the experimental hut trial. The insecticides evaluated were pyrethroids (transfluthrin 2 µg/bottle, alpha-cypermethrin 0.05%, permethrin 0.75%, deltamethrin 0.05%), a carbamate (bendiocarb 0.1%), an organochlorine (DDT 4%), and organophosphates (pirimiphos-methyl 170 mg/m²; malathion 5%). Transfluthrin (2ug/bottle) was tested in bottle bioassays while all other insecticides were tested in tube tests. For each insecticide, ∼100 females aged 3–5 days were tested in groups of 25 and exposed for 1 hour. Results were compared with the reference susceptible *An. gambiae* Kisumu strain. Knockdown was recorded after 1 h and mortality after 24 h.

### Data analysis

Count outcomes (numbers landing and numbers blood-fed) were pooled across both trial rounds and analyzed using negative binomial regression while proportional outcomes (mortality, blood-feeding, exophily) were analyzed using logistic regression models. Treatment arm was the primary fixed effect. Hut, trial round and day were also included as fixed effects, and sleeper was fitted as a random effect to account for individual variability and improve model precision.

Post-hoc power analysis: Using 1,000 simulations of a negative binomial regression model in Stata, we estimated the power to detect reductions in (i) blood-fed mosquitoes (aspiration, personal protection) and (ii) total mosquitoes collected (HLC, protective efficacy). Simulations incorporated observed IRRs, variance from model covariates, random effects for sleeper, and the overdispersion parameter. In both methods, all simulated *p*-values were <0.05, indicating 100% power.

### Ethical considerations

The trial was approved by the Ethics Review Committee of the Ministry of Health, Benin (CNERS Avis Éthique No. 18, 2024). Written informed consent was obtained from all participants after the study information sheet and consent form were thoroughly explained in the local language, with interpreters and impartial witnesses present where required. To minimize risk, only adult males aged 18–40 years were recruited, with women, children, and elderly individuals excluded. All participants were offered a free course of malaria chemoprophylaxis. A standby nurse was available throughout to monitor and manage any fever or adverse events. All methods were performed in accordance with the relevant guidelines and regulations.

## Results

### Susceptibility Bioassays

Mortality of pyrethroid-resistant *An. gambiae* s.l. from Covè was 100% following exposure to malathion, and bendiocarb in WHO tube tests, confirming full susceptibility to organophosphate and carbamate insecticides (Figure 2). In contrast, high levels of resistance were observed to pyrethroids and organochlorines, with <55% mortality to permethrin, deltamethrin, and alpha-cypermethrin, and <15% mortality to DDT (Figure 2). In transfluthrin (2 µg) bottle bioassays, knockdown was 100% but mortality only 32%, also confirming a high frequency of resistance to this insecticide. The susceptible *An. gambiae* Kisumu strain showed 100% mortality to all insecticides tested.

**Figure 2.**
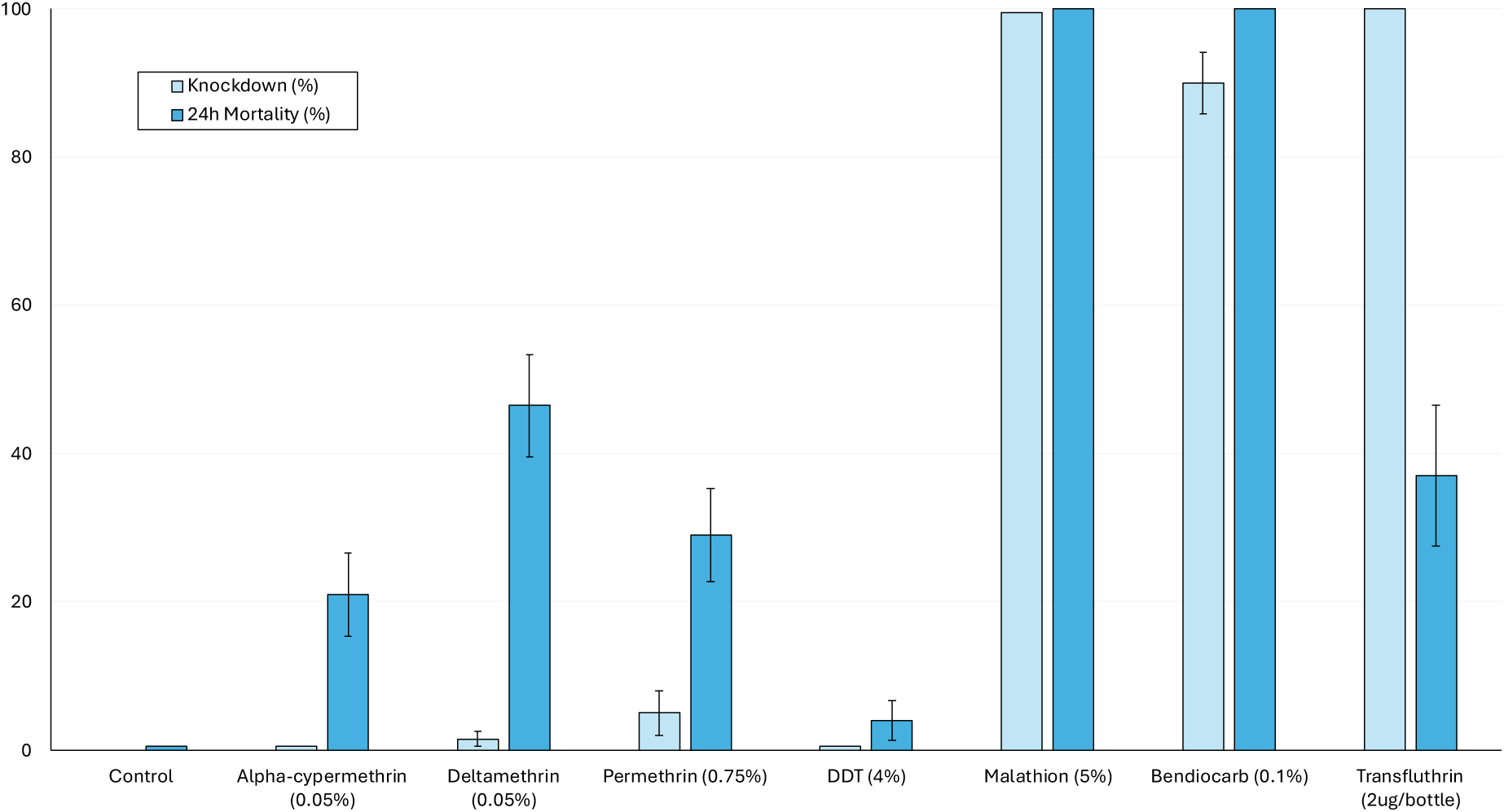
Knockdown and mortality of F1 progeny pyrethroid-resistant *Anopheles gambiae* s.l. (Covè strain) in WHO susceptibility bioassays conducted during the experimental hut trial. Transfluthrin was evaluated using bottle bioassays, while all other insecticides were assessed with WHO tube tests. The dashed red line represents the 98% threshold for susceptibility.

### Experimental hut trial results

A total of 5,682 wild, free-flying, pyrethroid-resistant female *Anopheles gambiae s.l.* were collected from the experimental huts across both trial rounds, comprising 4,246 from human landing catches (HLC) and 1,436 from mosquito aspiration collections. In addition, 6,158 *Mansonia africana*, a nuisance mosquito and vector of lymphatic filariasis, were collected by HLC. Although *Mansonia* were also present in aspiration catches, only specimens from the HLC collections were processed and included in the analysis due to time and resource constraints. Results from both trial rounds are presented in Table 1 and Figures 3–5 for HLC collections, and in Tables 2 and 3 for mosquito aspirations.

**Figure 3.**
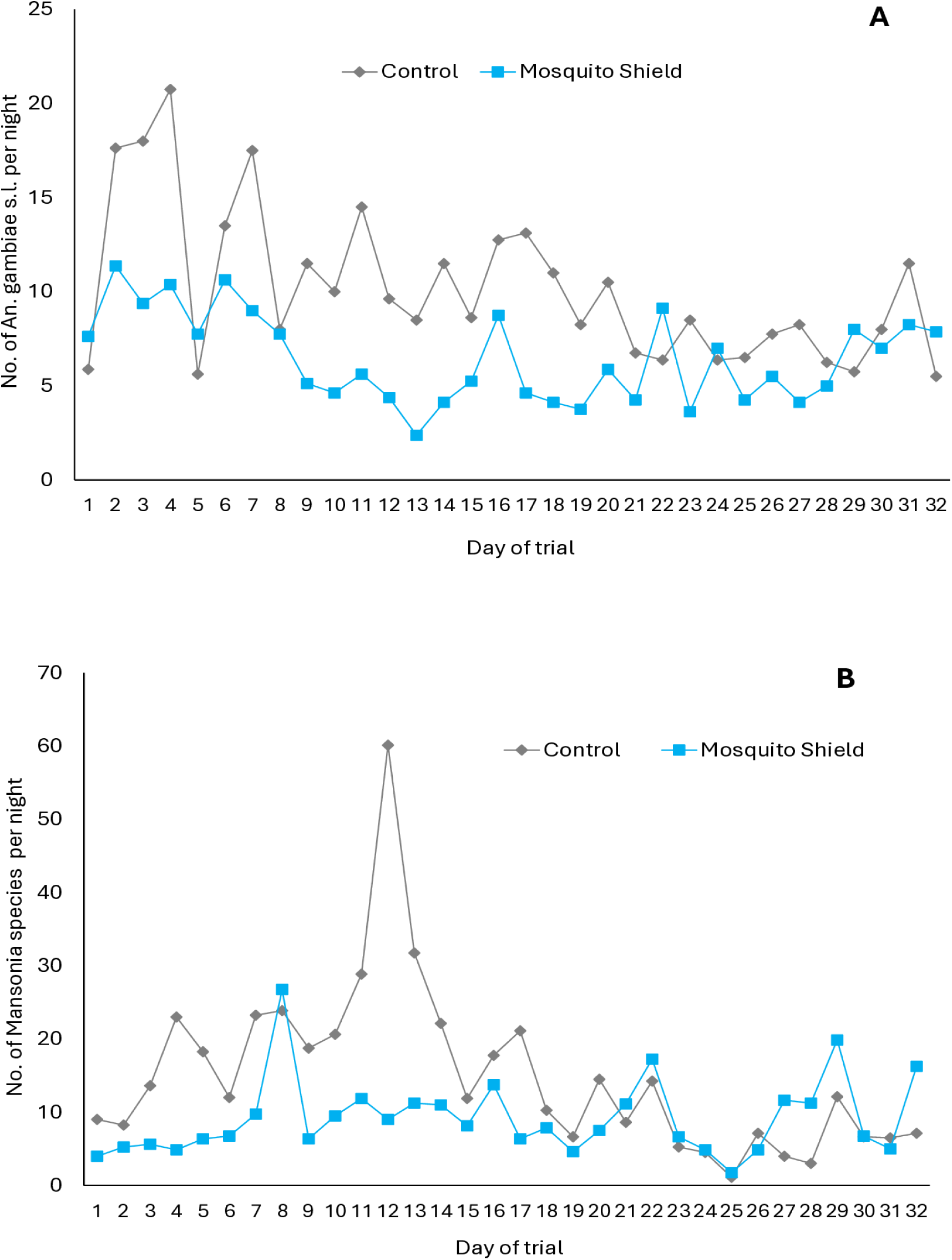
Human landing rates for pyrethroid-resistant *Anopheles gambiae* s.l. (A) and *Mansonia africana* (B) per study night in Mosquito Shield treated vs control huts. Data is pooled across both trial rounds.

**Figure 4.**
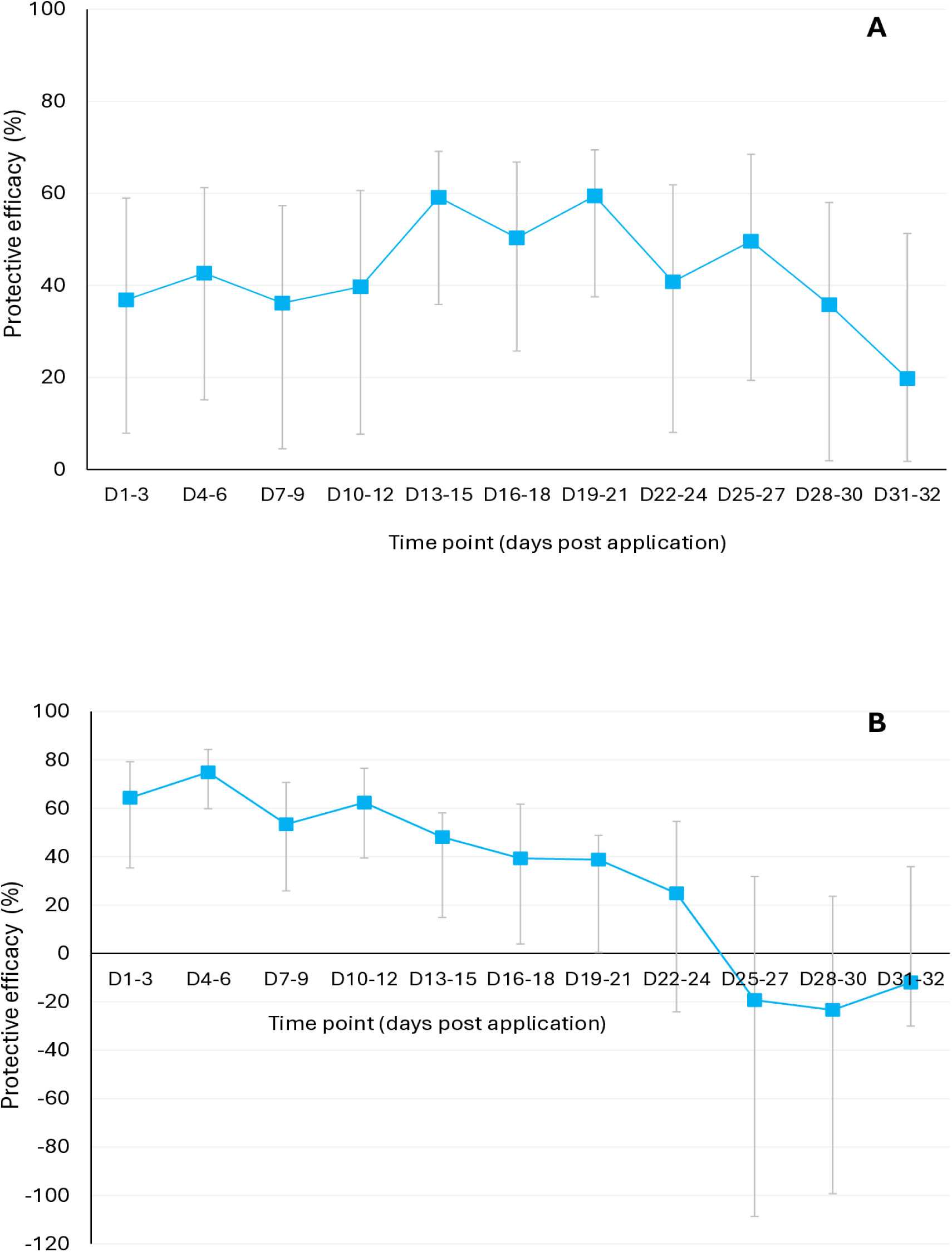
Protective efficacy of Mosquito Shield^TM^ against wild free-flying pyrethroid-resistant *Anopheles gambiae* s.l. (A) and Mansonia Africana (B) in experimental huts. Mosquitoes were collected by HLCs. Data is pooled across both trial rounds. D1 to D32 indicate the days of the trial post application of the intervention.

**Figure 5.**
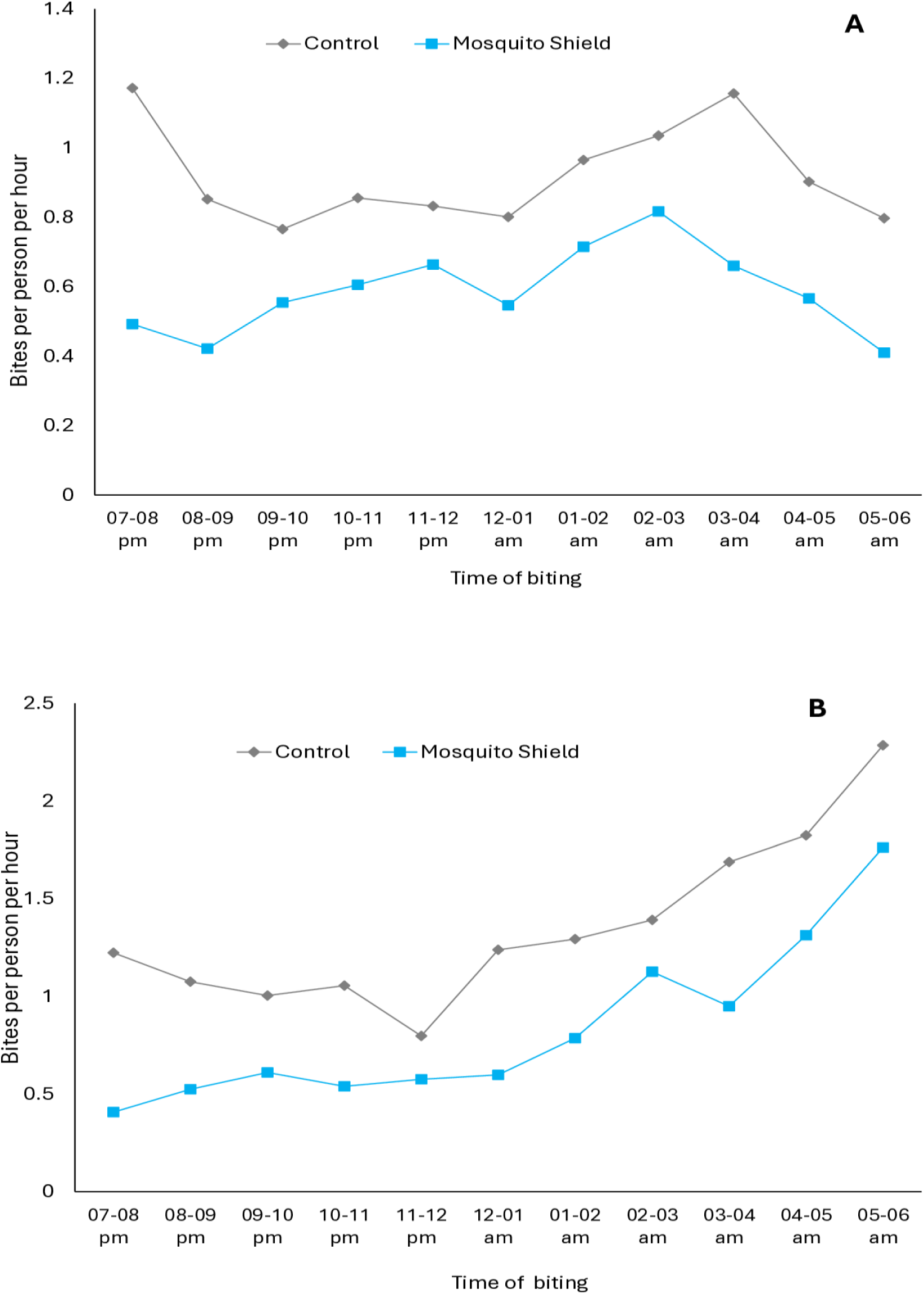
Hourly human landing rates per hour of wild pyrethroid-resistant *Anopheles gambiae* s.l. (A) and Mansonia africana (B) in Mosquito Shield treated vs control huts. Data is pooled across both trial rounds.

**Table 1.**
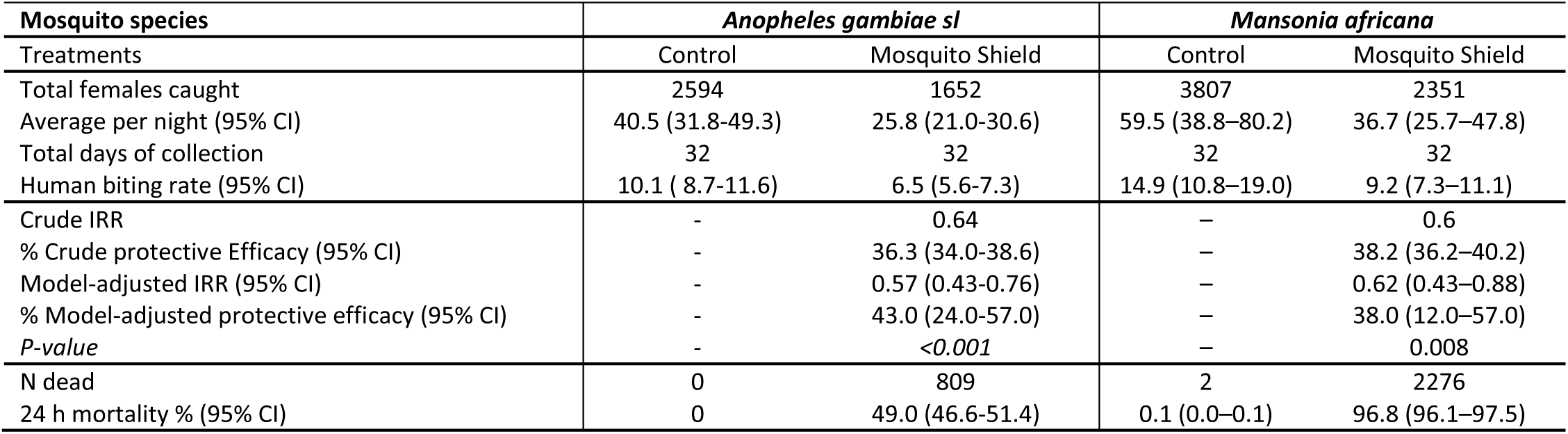
Human landing rates of wild mosquitoes in Mosquito Shield^TM^ treated experimental huts in Cove Benin (HLC method)

**Table 2:** Detterence, exiting and mortality of wild free-flying pyrethroid-resistant *An gambiae* s.l. in experimental huts in Cove, Benin. Mosquitoes were collected by aspirations performed early in the morning by volunteer sleepers.

| Treatments | Control | Mosquito Shield |
| --- | --- | --- |
| Total females caught | 948 | 488 |
| Average per night (95% CI) | 14.8 ( 10.8-18.8) | 7.6 (6.0-9.2) |
| Deterrence (%) | - | **48.5 (44.1-53.0)** |
| P-value | - | <0.001 |
| N exiting | 447 | 228 |
| Exophily (%) | **47.2 (44.0-50.3)** | **46.7 42.3-51.2)** |
| P-value | - | 0.962 |
| N inside net | 306 | 165 |
| % inside net | 32.3 | 33.8 |
| P-value | - | 0.607 |
| Total Dead | 0 | 111 |
| % Dead (95% CI) | 0.0 | 22.7 (19.0-26.5) |

**Table 3:** Personal protection against wild free-flying pyrethroid-resistant *An gambiae* s.l. in experimental huts in Cove, Benin. Mosquitoes were collected by aspirations performed early in the morning by volunteer sleepers.

| Treatments | Control | Mosquito Shield |
| --- | --- | --- |
| Total females caught | 948 | 488 |
| Total blood-fed | 305 | 110 |
| Crude IRR | - | 0.36 |
| Model-adjusted IRR (95% CI) | - | 0.37 (0.18-0.72) |
| **% Crude personal protection (95% CI)** | - | **63.9 (59.7-68.2)** |
| **% Model-adjusted personal protection (95% CI)** | - | **64.0 (21.9-81.8)** |
| P-value | - | 0.003 |
| Blood-fed (%) | 32.2 (29.2-35.1) | 22.5 (18.8-26.2) |
| P-value | - | <0.001 |
| Blood-feeding inhibition (%) | - | 29.9 (25.9-34.0) |

### Results with human landing catches

#### Wild free-flying pyrethroid resistant *Anopheles gambiae sl*

The number of wild free-flying pyrethroid resistant *Anopheles gambiae s.l* mosquitoes caught by HLC was substantially lower in huts treated with Mosquito Shield^TM^ compared to control huts (1,652 vs 2,594 females), corresponding to an average of 25.8 (95% CI: 21.0–30.6) versus 40.5 (95% CI: 31.8–49.3) mosquitoes per night (Table 1). The human biting rate was also reduced from 10.1 (95% CI: 8.7–11.6) bites per person per night in control huts to 6.5 (95% CI: 5.6–7.3) in treated huts. This represented a crude protective efficacy of 36.3% (95% CI: 34.0–38.6%) and a model-adjusted efficacy of 43.0% (95% CI: 24.0–57.0; p < 0.001), confirming a significant reduction in mosquito landing. Mortality among *An. gambiae s.l.* collected from Mosquito Shield^TM^ huts reached 49.0% (95% CI: 46.6–51.4%) compared to 0% in controls, demonstrating substantial insecticidal effects of the intervention.

Nightly catches of *Anopheles gambiae s.l.* were consistently lower in huts treated with Mosquito Shield^TM^ compared to untreated controls throughout the trial (Figure 3A). In control huts, catches often exceeded 10 mosquitoes per night, peaking above 20, whereas Mosquito Shield^TM^ huts rarely exceeded 10 and typically ranged between 3 and 7 per night. Protective efficacy against *An. gambiae s.l.* ranged between 30–60% during the 32-day evaluation period, with the highest levels observed between days 13 and 19, before declining to around 20% by day 32 (Figure 4A). Hourly biting activity was also reduced in Mosquito Shield^TM^ huts, where rates remained between 0.4 and 0.8 bites per person per hour compared to 0.8–1.2 in control huts, with the greatest reductions observed during peak evening (07:00–08:00 pm) and early-morning (03:00–04:00 am) biting periods, effectively halving exposure throughout the night (Figure 5A).

#### Wild free-flying *Mansonia africana*

For *Mansonia africana*, mosquito densities were substantially lower in huts treated with Mosquito Shield^TM^ compared to control huts, with 2,351 females collected versus 3,807 in controls (Table 1). The average nightly catch declined from 59.5 (95% CI: 38.8–80.2) in control huts to 36.7 (95% CI: 25.7–47.8) in treated huts, corresponding to a crude protective efficacy of 38.2% (95% CI: 36.2–40.2%) and a model-adjusted protective efficacy of 38.0% (95% CI: 12.0–57.0; p = 0.008). The human biting rate was similarly reduced, from 14.9 (95% CI: 10.8–19.0) bites per person per night in control huts to 9.2 (95% CI: 7.3–11.1) in treated huts. Mortality among *Mansonia africana* collected from Mosquito Shield^TM^ huts was exceptionally high at 96.8% (95% CI: 96.1–97.5%) compared to only 0.1% (95% CI: 0.0–0.1%) in controls, indicating strong and sustained lethality of the intervention against this species.

Nightly catches of *Mansonia africana* were consistently lower in huts treated with Mosquito Shield^TM^ compared to untreated controls throughout the trial (Figure 3B). In control huts, catches frequently exceeded 20 mosquitoes per night and peaked above 60 on day 12, whereas in Mosquito Shield^TM^ huts, counts remained relatively low, typically between 5 and 10 per night. Protective efficacy of Mosquito Shield^TM^ against *Mansonia africana* was initially high, exceeding 60% during the first two weeks post-application, but progressively declined over time, falling below zero by days 25–30, indicating a loss of effectiveness toward the end of the 32-day evaluation period (Figure 4B). Hourly biting activity of *Mansonia africana* increased steadily through the night, peaking between 05:00–06:00 am, with consistently lower biting rates in Mosquito Shield^TM^ huts (0.4–1.8 bites/person/hour) compared to controls (0.8–2.4 bites/person/hour), indicating substantial suppression of biting throughout all hours of the night.

### Results with mosquito aspirations

Across both trial rounds, 1,436 female *An. gambiae* s.l. were collected by the mosquito aspiration method, with 948 in control huts and 488 in Mosquito Shield^TM^ huts, corresponding to a 48.5% deterrence (95% CI: 44.1–53.0, p<0.001, Table 2). The average nightly catch was 14.8 mosquitoes (95% CI: 10.8–18.8) in control huts versus 7.6 (95% CI: 6.0–9.2) in Mosquito Shield huts. Exophily rates were similar between treatments, with 47.2% (95% CI: 44.0–50.3) in control huts and 46.7% (95% CI: 42.3–51.2) in Mosquito Shield^TM^ huts (p = 0.962). The proportion of mosquitoes found inside nets was also comparable, 32.3% in control versus 33.8% in Mosquito Shield^TM^ (p = 0.607)), indicating no consistent effect of Mosquito Shield^TM^ on mosquito entry inside the human occupied untreated net, although some variation was observed between the two trial rounds (Table S2). No mosquitoes died in the control huts, whereas 22.7% (95% CI: 19.0–26.5) died in Mosquito Shield^TM^ huts, indicating an insecticidal effect of the intervention.

The number of blood-fed mosquitoes was significantly lower in Mosquito Shield^TM^ huts (110) compared with controls (305, *aIRR*=0.03, p<0.001, Table 3). The proportion blood-fed was also reduced (22.5% vs. 32.2%), representing an overall blood-feeding inhibition of 29.9% (95% CI: 25.9–34.0, *p* < 0.001). Crude estimates of personal protection were 63.9% (95% CI: 59.7–68.2). Model-adjusted analyses showed even higher protective effects, with personal protection of 97% overall (*p* < 0.05). Personal protection levels against blood-feeding remained positive throughout the 32-day period (Figure 6). Protection peaked at about 81% on days 10–12, dipped to around 30% on days 19–21, and rose again to over 80% by days 31–32. These results confirm that Mosquito Shield^TM^ significantly and consistently reduced mosquito blood-feeding.

**Figure 6.**
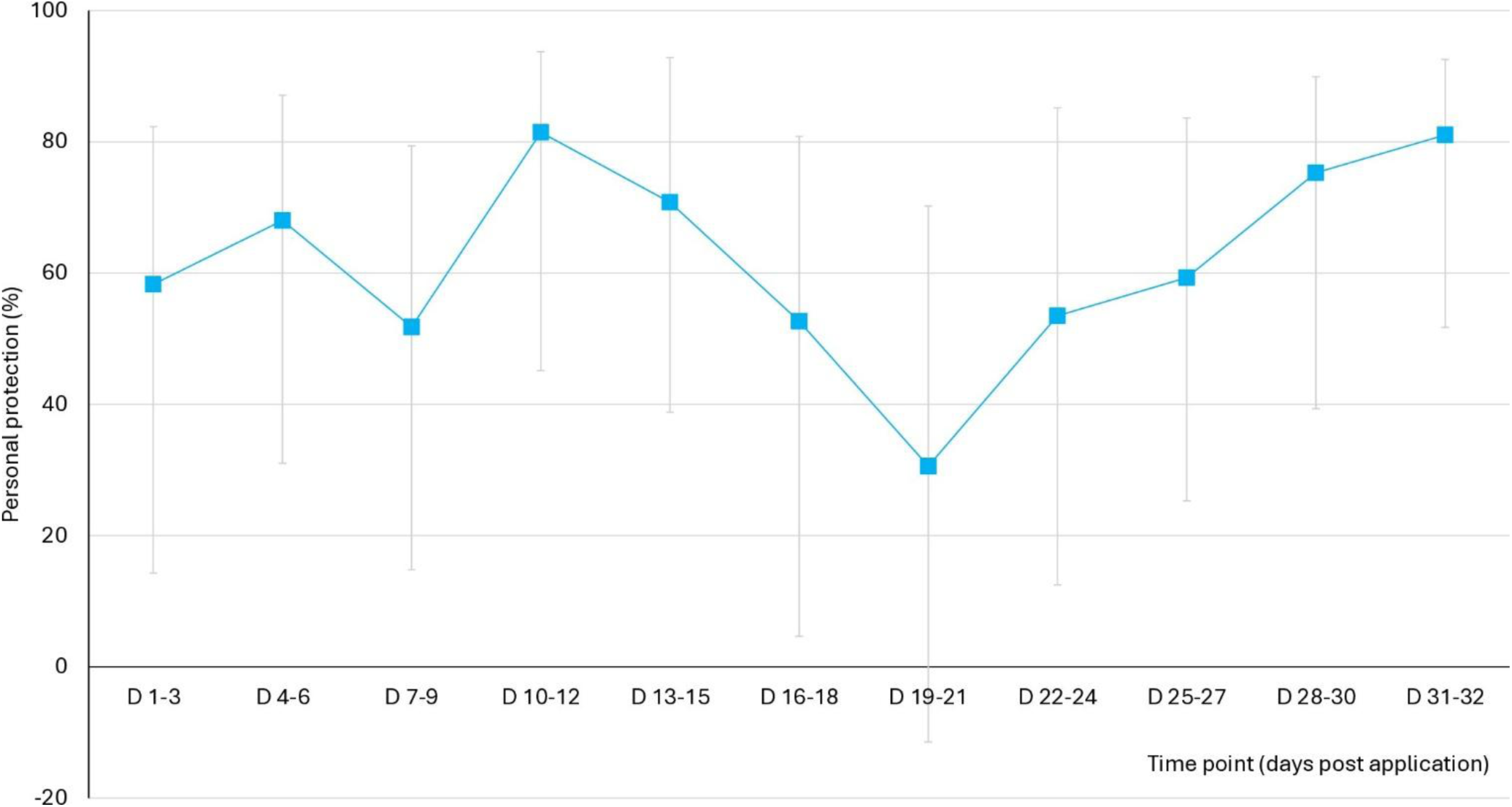
Personal protection of Mosquito Shield^TM^ against wild free-flying pyrethroid-resistant *Anopheles gambiae* s.l. in experimental huts. Mosquitoes were collected by aspirations. Data is pooled across both trial rounds. D1 to D32 indicate the days of the trial post application of the intervention.

## Discussion

This GLP-compliant trial provides evidence of the efficacy of Mosquito Shield–, a transfluthrin-based passive spatial emanator, in reducing human–vector contact under experimental hut conditions in West Africa, where high-intensity pyrethroid resistance is well established. Across two 32-day product life cycles and with complementary mosquito collection methods, the device consistently reduced *Anopheles gambiae s.l.* hut entry and human landing rates, significantly decreased blood-feeding, and lowered mosquito survival, resulting in substantial overall protective effects. These findings strengthen the growing evidence that passive emanator spatial repellents can be a valuable tool for malaria prevention, even in settings with intense pyrethroid resistance.

Across two transmission seasons, Mosquito Shield^TM^ reduced landing rates of wild, pyrethroid-resistant *An. gambiae* s.l., achieving 43% protective efficacy, consistent with a previous household randomized trial in Benin that reported 34.2% efficacy [8]. It also significantly reduced mosquito entry and blood-feeding, providing 30% blood-feeding inhibition and 64% personal protection. Although landing inhibition was lower than that observed in East African hut trials with less pyrethroid-resistant populations [12], these results are especially important for West Africa, where high-intensity pyrethroid resistance undermines ITN effectiveness [13, 14]. The findings show that Mosquito Shield^TM^ can substantially reduce mosquito biting and blood-feeding, reinforcing its potential as a complementary malaria prevention tool. Hourly biting profiles revealed the greatest impact during evening and early-morning peaks, when householders are least likely to be under nets [15]. This emphasizes the product’s capacity to close ITN protection gaps during periods of non-use, corroborating findings from community randomized trials [3] and supporting WHO’s recommendation for using spatial emanators alongside ITNs [6].

In addition to reducing mosquito landing and biting, Mosquito Shield^TM^ induced measurable mortality of wild *An. gambiae* s.l. in huts, despite the high levels of pyrethroid and transfluthrin resistance observed in WHO susceptibility bioassays. No mortality occurred in controls, whereas treated huts recorded ∼23–49% mortality depending on the mosquito collection method used. While mortality is not usually considered the primary mode of action of spatial emanators [16], these effects could still contribute to public health impact by removing a proportion of host-seeking vectors. Remarkably, the mortality levels observed were comparable to those achieved in standard WHO experimental hut trials of pyrethroid-only and pyrethroid-PBO nets against this vector population [17–19].

Nightly time-series data showed sustained suppression of mosquito entry and biting across the 32-day evaluation, with peak efficacy during the middle of the product cycle (days 13–19). A comparable mid-cycle performance pattern was observed in a previous household randomized trial in Benin [8], underscoring the consistency of these results. This indicates that the product provides optimal protection after an initial stabilization phase, with gradual decline toward the end of its active duration. These dynamics are critical for real-world deployment, underscoring both the reliability of protection over several weeks and the need to account for waning efficacy when planning replacement in areas with long transmission seasons. To address this, a longer-lasting spatial emanators with efficacy of up to 12 months has been developed and is currently under evaluation in Benin. Such a product would require fewer replacements and could be more cost-effective than Mosquito Shield^TM^ in settings with prolonged transmission periods.

The trial also recorded substantial effects against *Mansonia africana*, a nuisance mosquito and vector of lymphatic filariasis in West Africa [20]. Mosquito Shield^TM^ reduced *Mansonia* landing rates by 38% overall and achieved near-complete 24-h mortality (≥96%) across both rounds. Although the study was not designed to assess impact on this species, the results suggest potential co-benefits for integrated vector-borne disease control. Evidence from other studies shows that transfluthrin spatial emanators can also reduce transmission of arboviral diseases [21]. From a programmatic perspective, a single indoor device providing month-long protection against multiple vectors may be especially valuable in peri-urban settings where nuisance mosquitoes and arboviral vectors are abundant.

The study used two independent sampling methods in experimental huts, HLCs and mosquito aspirations, to evaluate the impact of Mosquito Shield^TM^ on malaria vectors, with landing rates and blood-feeding protection as the primary endpoints. HLCs are often regarded as the gold standard for assessing vector control tools, particularly for spatial emanators [16, 22, 23], but they are physically demanding for volunteers and raise ethical concerns [24]. Our results show that the standard WHO hut trial method based on mosquito aspirations can also generate robust endpoints under controlled semi-field conditions, offering a suitable high-throughput alternative for evaluating spatial emanators. This method provides a more realistic representation of the interactions of the product with humans when applied indoors and allows assessment of a broader range of entomological outcomes, including deterrence, exophily, mortality, personal protection, blood-feeding inhibition, and proportion inside nets, providing a more comprehensive evaluation of product performance. These findings align with recent WHO testing guidelines for spatial emanators, which recommend incorporating additional entomological endpoints beyond human landing rates [25].

In conclusion, Mosquito Shield^TM^ reduced entry, landing, blood-feeding, and survival of *An. gambiae* s.l. and strongly suppressed *Mansonia africana* under semi-field experimental hut conditions in an area of high pyrethroid resistance. While its public health impact in West Africa remains to be demonstrated, the results underscore its potential to strengthen malaria control and provide additional benefits for integrated vector-borne disease management in the region. Importantly, these findings supported WHO’s prequalification of Mosquito Shield–[6, 26], paving the way for broader adoption and incorporation into national vector control strategies.

## List of abbreviations

ITN: Insecticide treated nets
IRS: Indoor residual spraying
WHO: World Health Organization
AI: Active ingredient
HLC: Human landing catches
HBR: Human biting rate
IIR: Incidence Rate Ratio
aIIR: adjusted Incidence Rate Ratio
RCT: Randomised controlled trials
CREC: Centre de Recherche Entomologique de Cotonou
LSTM: Liverpool School of Tropical Medicine
PAMVERC: Pan African Malaria Vector Research Consortium
AIRID: African Institute for Research in Infectious Diseases

## Declarations

### Availability of data and material

The datasets generated and analysed during the current study contain information from human volunteers and are subject to ethical and regulatory restrictions. De-identified data sufficient to reproduce the analyses are available from the corresponding author upon reasonable request.

### Consent for publication

Not applicable

### Authors’ contributions

CN acquired funding, designed the study and supervised its implementation. BN, JA and MY performed the hut trial with support from TS. TS, BN and CN analysed the data and prepared the manuscript tables and figures. CN and BN wrote the manuscript text with support from TS, BP and AY. All authors read and approved the final manuscript.

## Acknowledgements

We thank Dr. Thomas Mascari and Mrs. Madeleine Conaway of SC Johnson & Son, Inc. for providing the test items and their valuable support. We are grateful to the rice farmers of Cové for their participation and collaboration, and to the staff of the CREC/PAMVERC/AIRID Benin Research Programme (including Imelda Glele, Apithy Danielle, Nadia Houeto, Damien Todjinou, and others) for their dedicated assistance throughout the study.

